# The preoperative abundance of *Prevotella* associates with anastomotic healing after colorectal surgery

**DOI:** 10.1101/2021.10.25.21265372

**Authors:** Konstantina Zafeiropoulou, Emmeline G. Peters, Daan J. Brinkman, Andrew Y.F. Li Yim, Boudewijn J.J. Smeets, Theodorus B.M. Hakvoort, Evgeni Levin, Mark Davids, Misha D.P. Luyer, Wouter J. de Jonge, Joep P.M. Derikx

## Abstract

Anastomotic leakage (AL) is a frequent and potentially life-threatening postoperative complication in patients undergoing colorectal surgery with a multifactorial yet unknown pathophysiology. In the present study, we aimed to unravel AL etiology and intestinal anastomotic healing (AH) physiology focusing on both host epithelial and gut microbial components. We make use of patients-related samples and two high throughput next generation sequencing analysis approaches, including anastomotic tissue-associated bulk RNA sequencing and preoperative fecal 16S rRNA gene sequencing. We find that in the absence of host epithelial gene expression differences, the preoperative fecal microbiota of patients who undergo colorectal resection with successful AH differs in the group of patients who develop AL in the abundance of *Prevotella* and suggests a potential beneficial role of *Prevotella* that may prove useful as predictive biomarker for AH.

---

Anastomotic leakage (AL) is a severe postoperative complication in patients undergoing colorectal surgery, which negatively impacts hospital costs, morbidity, mortality, re-operation rates, and duration of hospitalization (Hammond et al., 2014). Although immunosuppressive medication and distal location of the anastomosis are known risk factors for AL (Sahami et al., 2016), its pathophysiology remains poorly understood and options to prevent AL are limited. The unpredictability of AL can be attributed to a lack of fundamental understanding of the processes underlying anastomotic healing (AH) (Bosmans et al., 2015). The current hypothesis suggests a multifactorial etiology, with host genetic make-up, perioperative inflammation and intestinal microbiota being key players (Lee et al., 2018).

Previous work from our group has demonstrated an association between AL and postoperative ileus (POI) (Peters et al., 2017). Although POI-associated stasis of the luminal content and preoperative microbiome signal is now profound (Shogan et al., 2020), the role of gut microbiota in AL remains unclear. Earlier studies point to low abundance of collagenase-producing strains (i.e. *Enterococcus faecalis*) that may predominate in the lumen of patients with colorectal cancer (CRCA) and drive AL via collagen breakdown (i.e., matrix metalloproteinase 9) (Shogan et al., 2015). Further findings show a predictive value of the anastomotic tissue-related mucus microbiome in the absence of C-seal treatment, characterized by decreased microbial diversity and increased relative abundance of mucin-degrading commensal members (van Praagh et al., 2019). However, there is currently no consensus on AL-associated gut microbial dysbiosis. Respectively, findings from the latter cohort indicate pre-existing differences in the gene expression, with the collagen cross linking pathway being downregulated (van Praagh et al., 2021), but that needs to get confirmed in larger cohorts and mechanistically challenged.

In the present study, we explored the AL pathogenesis in a stepwise approach using two high-throughput next generation sequencing techniques (i.e., tissue RNA sequencing and fecal 16S rRNA gene sequencing). Patients with CRCA who underwent colorectal resection were herein included; all patients were enrolled in a multicenter double-blind randomized controlled study from August 2014 until February 2017 (SANICS II trial) (Peters et al., 2018).

We initially interrogated the anastomotic tissue-associated transcriptome at time of surgery. Samples were obtained from a SANICS II-subgroup of 18 patients who developed AL after colorectal surgery, and 18 age, sex and body mass index (BMI)-matched patients with successful postoperative AH (Table 1). A pipeline of pre-processing steps was applied, including quality control of the RNA sequencing raw reads (Andrews, 2010; Ewels et al., 2016), alignment to the human genome (Dobin et al., 2013), annotation (Aken et al., 2017), post-alignment processing analysis (Li et al., 2009) and reads counting (Liao et al., 2013). Unsupervised exploratory analyses suggested no clear separation between AL and AH through hierarchical clustering analysis for the 150 most variable genes (Figure S1A). Subsequent supervised analyses (Robinson et al., 2010) identified no difference in expression observed between the AL and AH patient cohorts either (Figure S1B). This finding highlighted further our need to address the role of gut microbiota in the process of AL, rather than pre-existing differences in host gene expression at the time of surgery.

**Table 1.**
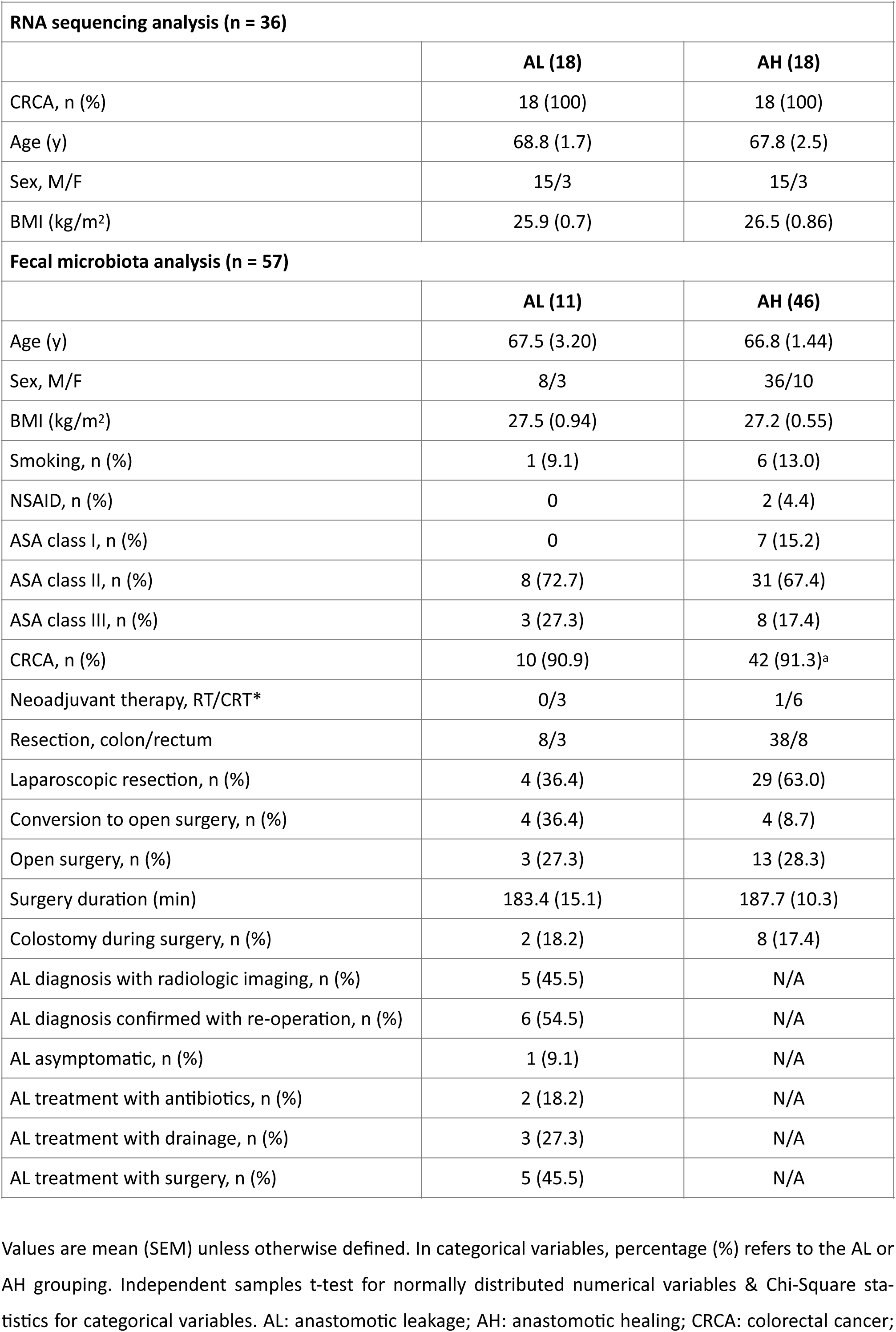
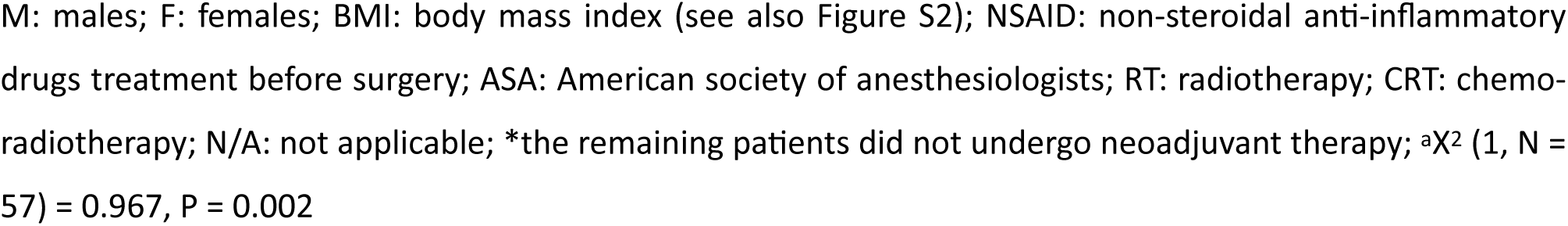
Patient Characteristics.

| **RNA sequencing analysis (n = 36)** |  |  |
| --- | --- | --- |
|  | **AL (18)** | **AH (18)** |
| CRCA, n (%) | 18 (100) | 18 (100) |
| Age (y) | 68.8 (1.7) | 67.8 (2.5) |
| Sex, M/F | 15/3 | 15/3 |
| BMI (kg/m<sup>2</sup>) | 25.9 (0.7) | 26.5 (0.86) |
| **Fecal microbiota analysis (n = 57)** |  |  |
|  | **AL (11)** | **AH (46)** |
| Age (y) | 67.5 (3.20) | 66.8 (1.44) |
| Sex, M/F | 8/3 | 36/10 |
| BMI (kg/m<sup>2</sup>) | 27.5 (0.94) | 27.2 (0.55) |
| Smoking, n (%) | 1 (9.1) | 6 (13.0) |
| NSAID, n (%) | 0 | 2 (4.4) |
| ASA class I, n (%) | 0 | 7 (15.2) |
| ASA class II, n (%) | 8 (72.7) | 31 (67.4) |
| ASA class III, n (%) | 3 (27.3) | 8 (17.4) |
| CRCA, n (%) | 10 (90.9) | 42 (91.3)<sup>a</sup> |
| Neoadjuvant therapy, RT/CRT* | 0/3 | 1/6 |
| Resection, colon/rectum | 8/3 | 38/8 |
| Laparoscopic resection, n (%) | 4 (36.4) | 29 (63.0) |
| Conversion to open surgery, n (%) | 4 (36.4) | 4 (8.7) |
| Open surgery, n (%) | 3 (27.3) | 13 (28.3) |
| Surgery duration (min) | 183.4 (15.1) | 187.7 (10.3) |
| Colostomy during surgery, n (%) | 2 (18.2) | 8 (17.4) |
| AL diagnosis with radiologic imaging, n (%) | 5 (45.5) | N/A |
| AL diagnosis confirmed with re-operation, n (%) | 6 (54.5) | N/A |
| AL asymptomatic, n (%) | 1 (9.1) | N/A |
| AL treatment with antibiotics, n (%) | 2 (18.2) | N/A |
| AL treatment with drainage, n (%) | 3 (27.3) | N/A |
| AL treatment with surgery, n (%) | 5 (45.5) | N/A |

Values are mean (SEM) unless otherwise defined. In categorical variables, percentage (%) refers to the AL or AH grouping. Independent samples t-test for normally distributed numerical variables & Chi-Square statistics for categorical variables. AL: anastomotic leakage; AH: anastomotic healing; CRCA: colorectal cancer;

Therefore, we included a SANICS II-subgroup of 57 patients (13 women) who participated in the preoperative fecal microbiota study providing a fecal sample one day prior to the colorectal surgery; 11 out of the 57 patients developed AL, while the remaining 46 did not (AH). AL was diagnosed based on radiologic imaging (5 out of 11 AL) or confirmed during re-operation (6 out 11 AL) and treated with antibiotics (2 out of 11), drainage (3 out of 11 AL), secondary surgery (5 out of 11 AL) or untreated in the absence of symptoms (1 out of 11 AL, Table 1). There was no association between AL development and resection’s location (46 colonic; 11 rectal resections), neoadjuvant therapy (chemo-radiotherapy in 3 out of 11 AL) or performance of colostomy (9 out of 57 patients); no difference in surgery duration between AL and AH was found either (Table 1). Thirty-three patients underwent a laparoscopy and 16 an open resection, while eight patients were converted to open surgery, which tended to be significantly associated with the development of AL (X^2^ (2, N = 57) = 5.936, P = 0.051, Table 1). There was no difference in age, sex nor BMI between the AL and AH patients, whilst no association was found between smoking behavior and postoperative diagnosis of AL. All 57 patients were classified up to American society of anesthesiologists (ASA) class III; two out of the 57 were treated with non-steroidal anti-inflammatory drugs (NSAID) before surgery, with no association between the ASA score or NSAID treatment and AL retrieved (P > 0.05 for all aforementioned analyses, Table 1).

We profiled the preoperative fecal microbiota structure of all 57 patients using 16R rRNA gene sequencing targeting the V3-V4 regions (Kozich et al., 2013). A pipeline of pre-processing steps was herein conducted as well, including reads truncation, merging, amplicon sequence inference (Edgar, 2016), and annotation using the SILVA 16S ribosomal database 132 (Quast et al., 2013); amplicon sequence variants (ASV) complete count table was stored as a Phyloseq object (McMurdie and Holmes, 2013). We found no difference in fecal α diversity between AL and AH groups, arguing against a profound dysbiosis pre-existing preoperatively (Figure 1A). AL development could not explain the variation in fecal ASV community structure (explored using the vegan package (Oksanen J., 2020), Figure 1B), which was further confirmed by the local contribution to β diversity (LCBD) analysis where no difference was observed between the two groups (Figure 1A). Using the Bioenv workflow, a subset of ASVs belonging to 21 genera was suggested as major determinants of the community complexity, explaining 83.5% of the variance described by the complete ASV dataset (Table S1).

**Figure 1.**
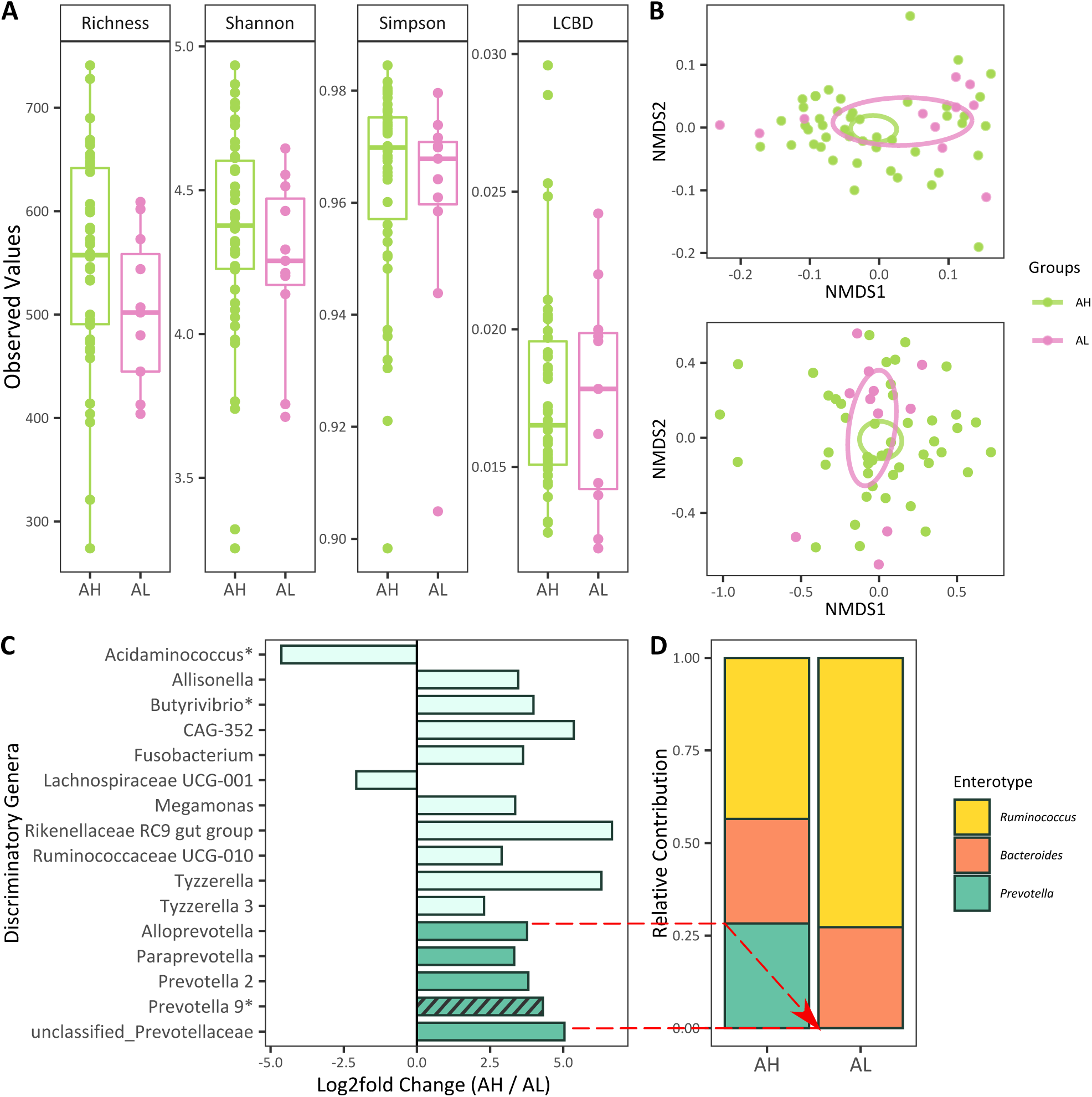
Preoperative abundance of Prevotella associates with AH after colorectal surgery in the absence of profound preoperative AL-related fecal microbial dysbiosis. (A) Alpha diversity indices (i.e., Richness, Shannon, Simpson diversity) and local contribution to beta diversity (LBCD) of preoperative fecal samples (rarefied to 30,000 reads) from patients who underwent colorectal surgery (n = 57); (B) Nonmetric multidimensional scaling plots of the fecal beta diversity based on unweighted UniFrac distance (upper panel) and Bray-Curtis dissimilarity index (lower panel); (C) Log2fold changes of the 16 discriminatory between AL and AH patients genera (P (adjusted) < 0.05, see also Table S2 and Figure S2); asterisk indicates Bioenv-selected taxa suggested as major determinants of the community complexity (see also Table S1); stripes indicate influential taxa based on the conventional random forest classification-feature importance analysis; taxa essential components of the Prevotella enterotype are indicated with dark green (same color as Prevotella enterotype in panel (D)); the red dashed arrows highlight the significant absence of Prevotella-related taxa in AL; (D) Relative contribution of the three enterotypes in the AL and AH patients.

We then looked at the preoperative fecal microbial composition pursuing differential analysis using the DESeq2 package (Love et al., 2014) in taxa relative abundance between patients who developed AL after surgery and those who did not at both ASVs and agglomerated at genus levels. All 57 patients were characterized by a total of 2,577 distinct, non-mitochondrial ASVs. One hundred and six ASVs out of the 2,577 (3.9%) differed significantly between the two groups, with 90 and 16 being significantly increased and decreased in feces of AH patients, respectively (Table S2). At genus level, 16 taxa differed significantly between AL and AH, with only two being significantly increased in patients who developed AL after surgery (P (adjusted) < 0.05, Figure 1C and Table S2). The remaining 14 displayed significantly increased preoperative relative abundance in the feces of AH group, suggesting a potential beneficial role in intestinal AH.

To challenge the meaning of these significant preoperative fecal microbial composition differences in the development of AL and physiology of AH, we explored potential correlations between their relative abundances, patient-related individual factors (i.e., age, BMI) and preoperative serum concentrations of the TNF-α, interleukin 1β (IL-1β), IL-6, IL-8, IL-10 and IL-12 cytokines. Of the 16 discriminatory genera, only the relative abundance of *Paraprevotella* correlated significantly negatively with the preoperative serum concentration of the cytokine IL-12, suggesting that the remaining 15 were less likely to be explained by differences in the inflammatory status between the two groups, but rather AL- and AH- specific (Figure S2).

*Prevotella 2, Prevotella 9, Alloprevotella, Paraprevotella* and unclassified *Prevotellaceae*, five out of the 14 (35.7%) discriminatory taxa with significantly increased relative abundance in the AH group, are all amongst the essential components of the *Prevotella* enterotype (highlighted with dark green in the bar plot in Figure 1C). In the present cohort, *Prevotella 9* was also listed among the 21 Bioenv-selected taxa found to explain 83.5% of the fecal microbial variance (indicated with asterisk in Figure 1C, Table S1), signifying its importance in the microbiome structure of the 57 patients. Therefore, we proceeded one step further and performed statistical analysis at enterotypes level (Arumugam et al., 2011) that highlighted the beneficial effect of preoperative *Prevotella* when no subjects with a predicted *Prevotella* enterotype developed AL (X^2^ (2, N = 57) = 7.658, P = 0.10, Figure 1D).

Our data suggest a beneficial role of *Prevotella* in AH. We confirmed it further with the means of feature importance analysis. Using the complete count table aggregated at genus level, conditional random forest classifier (Hothorn et al., 2006) distinguished patients who developed AL from patients who did not with an “out-of-bag” error rate of 19.3%. This was significantly more effective than random classification (permutation analysis of variance P = 0.019 and area under the curve = 0.660). Importantly, genera *Prevotella 9* and *Prevotella* 7 were among the most influential taxa, with conditional variance importance suggesting them as the third (variance importance = 4E-04) and twelfth (variance importance = 2E-04) most prominent genera respectively (indicated with stripes at Figure 1C).

Overall, we demonstrated that in the absence of host gene expression differences, the gut microbiota of patients who undergo colorectal resection with successful AH differs in the group of patients who develop AL in the abundance of *Prevotella*. We used an elaborative pipeline of statistical analyses that enables us to suggest that the *Prevotella*-related microbial signals in the group of patients who do not develop AL may prove useful as predictive biomarkers for AH. *Prevotella* enterotype has been distinctly described in loose feces, as defined by increased Bristol Stool Scale ratings of their stool consistency, a proxy for the intestinal colon transit time (Vandeputte et al., 2016). The abundance of *Prevotella* enterotype in subjects with loose feces, may render easier gastrointestinal passage resulting in less stasis and luminal distention and thus better anastomotic healing, a functional partial explanation for the present findings. We speculate that passage of harder stools will hinder an anastomosis from healing.

Decreased *Prevotella* has been also described in AL mucus-associated microbiome, when bacterial DNA was isolated from “donuts” of patients in whom a stapled colorectal anastomosis was made, yet that was not significant when the non C-seal and C-seal covered anastomosis patients were analyzed together (van Praagh *et al*., 2019). In following up analyses, we wish to validate a *Prevotella*-associated predictive algorithm for AH in a separate cohort of patients (i.e., REVEAL cohort (Jongen et al., 2016)). We believe that this can have a significant clinical and economic impact once implemented in CRCA surgeries, where the surgeon has options to prevent surgical anastomoses in patients at risk for developing AL.

## Data Availability

All data produced in the present study are available upon reasonable request to the authors.

## Acknowledgements

The authors would like to thank all SANICS II trial participants. J.D. and W.J are funded by the Dutch Research Council (ZonMW-NWO) research program Vidi project, No. 91719343 and AMC-stipendium 200402.

## Authors Contributions

Conceptualization, E.P., W.J. and M.L.; Data curation, E.P.; Formal analysis. K.Z., A.L.Y. and M.D., Funding acquisition: W.J., J.D. and M.L.; Investigation: E.P., D.B., B.S. and T.H.; Project administration: E.P.; Resources: W.J.; Supervision: E.L., J.D., M.L. and W.J.; Visualization: K.Z. and A.L.Y.; Writing – original draft: K.Z.; Writing – review & editing: K.Z., J.D. and W.J.

## Data and Code Availability

All data produced in the present study are available upon reasonable request to the authors.

### Additional Resources

The study was registered at ClinicalTrials.gov (NCT02175979, https://clinicaltrials.gov/ct2/show/NCT021759 79) and trialregister.nl (NTR4670, https://www.trialregister.nl/trial/4494).

### Ethical Considerations

The study was approved by the Medical Ethics Committee of the Catharina Hospital. Informed consent was obtained from all subjects.

## Supplemental methods

### Human subjects

All subjects included in the present study were recruited in the SANICSII trial, which was open for inclusion from August 2014 until February 2017, performed to compare perioperative nutrition to standard care with POI as primary endpoint. ASA physical status score was used to assess their fitness before surgery. C-reactive protein was measured to evaluate inflammation postoperatively with four consecutive daily measurements. AL was defined radiologically or confirmed during re-operation. Patients who developed AL were treated with antibiotics, drainage, secondary surgery or untreated in the absence of symptoms.

In total 280 patients were included, all of which had provided a written informed consent. The current study pertained a subset of 36 patients used to interrogate the anastomotic-tissue associated transcriptome at the time of surgery, and a subset of 57 patients used to evaluate the preoperative fecal microbiota. Information about all patients’ age, sex, BMI and postoperative complications (AL or AH) is provided in Table 1. Further detailed information about patients’ pre- (i.e. NSAID treatment, ASA class and neoadjuvant therapy), postoperative (i.e., AL diagnosis and treatment) metadata, and surgery’s characteristics (i.e., colonic or rectal resection, laparoscopic, open surgery or conversion to open surgery, and formation of colostomy during surgery) are also reported in Table 1; as no significant difference in the host transcriptome between AL and AH patients, detailed information about pre-, peri- and postoperative metadata are only reported for the 57 patients included in the fecal microbiota cohort.

### RNA sequencing

RNA was isolated from human full thickness colonic tissue using the Isolate II RNA mini kit (Cat# BIO-52073, Meridian Bioscience). RNA quality was assured using the Bioanalyzer (Agilent, Santa Clara, USA), including only samples with a RIN score higher than 8. Conversion of the mRNA to cDNA was done using the NEBNext Ultra Directional RNA Library Prep Kit (Cat# E7760L, New England Biolabs) for Illumina and subsequent sequencing of the cDNA was performed on the NextSeq500 to a depth of 10 M reads per sample. Both cDNA conversion and sequencing were performed at GenomeScan (Leiden, The Netherlands).

Quality control of the raw reads was done with FastQC (version 0.11.5) (Andrews, 2010) and the summarization thereof through MultiQC (version 0.8) (Ewels *et al*., 2016). Raw reads were aligned to the human genome (GRCh38) using STAR (version 2.5.3a) (Dobin *et al*., 2013) and annotated using the Ensembl v91 annotation (Aken *et al*., 2017). Post-alignment processing was performed through SAMtools (version 1.2) (Li *et al*., 2009), after which reads were counted using the featureCounts application in the Subread package (version 1.6.0) (Liao *et al*., 2013).

### Fecal sample collection

Feces were collected by the patients on the day prior to surgery and immediately stored at -20°C.

### Microbiota profiling

DNA was extracted from 150 mg of fecal sample using mechanical lysis and the PSP Spin Stool DNA Plus Kit (Cat# 1038110300, STRATEC Molecular GmbH) isolation method. The samples were placed in Lysing Matrix E tubes with 1,400 ul of stool stabilizer from the PSP kit. Mechanical lysis was done using the FastPrep at 3 repetitive rounds of 30 sec at 6.5 m/s, with cooling of 30 sec on ice in between. After the FastPrep procedure further extraction was done at 95°C for 15 min followed by cooling on ice for 1 min and centrifugation at 13,400 g for 1 min. The supernatant was transferred to the PSP InviAdsorb. Negative extraction controls (DNA-free water) were processed in the same way. The DNA concentration was measured with the Nanodrop 1000 spectrophotometer (Thermo Fisher Scientific).

16S rRNA gene amplicons were generated using a single step PCR protocol targeting the V3-V4 region (Kozich *et al*., 2013); both forward and reverse primers are listed in the key resources table. The PCR reaction in 20 ul was performed with following thermocycling conditions: initial denaturation at 98°C followed by 25 cycles denaturation (10 sec at 98°C), annealing (20 sec at 55°C) and extension (90 sec at 72°C) and final extension at 72°C for 10 min. PCR products were purified using Ampure XP beads (Cat# A63882, Beckman Coulter Life Sciences) and purified products were equimolar pooled. The libraries were sequenced using a MiSeq platform using V3 chemistry with 2×251 cycles.

Forward and reverse reads were truncated to 240 and 210 bases respectively and merged using USEARCH; merged reads that did not pass the Illumina chastity filter, had an expected error rate higher than 2, or were shorter than 380 bases were filtered. Amplified sequence variants (ASVs) were inferred for each sample individually with a minimum abundance of 4 reads (Edgar, 2016). Unfiltered reads were then mapped against the collective ASV set to determine the abundances. Taxonomy was assigned using SILVA 16S ribosomal database V132 (Quast *et al*., 2013). Enterotype classification was done based on the Jensen-Shannon divergence-clustering of the samples, as published by the MetaHIT consortium in 2011 https://enterotype.embl.de/enterotypes.html.

### Serum cytokines measurement

Blood samples were collected from all patients the day before surgery. They were immediately put on ice, centrifuged at 4°C and 3,000 g for 12 min and the plasma was stored at − 80°C until further analysis. To determine the most important inflammatory cytokines, a Human Inflammatory Cytokines Kit was used to measure cytokine levels of IL-1β, IL-6, IL-8, IL-10, IL-12, and TNF-α by cytometric bead array (Cat# 551811, BD Biosciences). Standards of 9 distinct concentrations were used (i.e., 5,000, 25,000, 1,250, 625, 312.5, 156, 80, 40 and 20 pg/mL), along with a blank sample. Mix bead solution was prepared and incubated with PE detection reagent and the samples for 3 hrs, at room temperature, in 96-well plate V bottom wells. Samples were measured on a BD LSRFortessa (BD, Biosciences) and analyzed using BD Flowjo Software (version 10.7.1).

### Quantification and statistical analysis

Differential expression analysis of the RNA sequencing data was performed using edgeR (version 3.20.6) (Robinson *et al*., 2010) in the R statistical environment (version 3.4.3). A quasi-likelihood F-test (QL F-test) was performed using the glmQLFit function to find differentially expressed genes using the estimated trended dispersions; results presented in Figure S1. Regressions were corrected for age and sex.

Clinical metadata were analyzed using IBM SPSS Statistics for Windows (version 26.0); both one-sample Kolmogorov-Smirnov and Shapiro-Wilk tests were used to confirm normal distribution in numerical variables. Chi-Square test of Independence was applied to explore associations between categorical variables. Parametric independent-samples t-test was conducted to compare the means of normally distributed variables between the groups. All statistical parameters are reported in Table 1; in the categorical variables, n represents the exact number of patients and percentage (%) refers to the respective AL or AH grouping. In the numerical variables, values present the mean, and standard error of the mean follows within brackets.

Microbial multivariate statistical analysis was performed in R statistical environment (version 4.3.0) using the packages phyloseq (version 1.34.0) (McMurdie and Holmes, 2013) (Oksanen J., 2020) and vegan (version 2.5.7). Samples were rarefied to 30,000 reads. Alpha diversity indices were calculated (i.e., species richness, Shannon entropy, Simpson diversity index). Microbial composition was assessed using non-metric multidimensional scaling plots (NMDS) at ASV level based on Bray-Curtis dissimilarity index and unweighted UniFrac distance. The former considers bacterial taxon abundance, while the latter considers phylogenetic distance between bacterial taxa through presence/absence, regardless of proportional representation. Non-parametric permutational multivariate analysis of variance (PERMANOVA) was applied using the vegan Adonis function on distance matrices (i.e., Bray-Curtis dissimilarity index and unweighted UniFrac distance). Local contribution to β diversity (LCBD) analysis was performed to measure the contribution of each sample to the total ASV β diversity; samples with high LCBD represent samples that are markedly different from the average β diversity of all study samples. Alpha and beta diversity results are presented in Figure 1A and 1B. Differences in ASV and genus relative abundances between groups were found using the DESeq2 method (version 1.30.1) (Love *et al*., 2014). Basemean, log2-fold change, non-adjusted and adjusted p-values of the aforementioned analyses at ASV and genus levels are reported in Table S2. For correlations Kendall rank correlation was used; results presented in Figure S2. Benjamini-Hochberg correction was applied to cases of multiple testing. Analysis using the Bioenv function in vegan produced subsets of taxa whose Euclidean distance matrices correlate maximally with the Bray-Curtis dissimilarity matrices derived from complete count tables, thus indicating major determinants of community structure; results presented in Table S1. Conventional random forest analysis used the complete count table aggregated at genus level. Models were generated using Log-proportional abundances via the cforest function of the party R package (version 1.3.7), which creates random forests from unbiased classification trees based on a conditional inference framework (Hothorn *et al*., 2006). Feature importances were evaluated calculating the conditional variance importance; respective results highlighted on Figure 1C.

**Table S1.** Bioenv selected taxa explain 83.5% of the variance in fecal microbiota structure of preoperative patients described by the complete ASV dataset agglomerated at genus level.

|  |
| --- |
| *Faecalibacterium* |
| *Subdoligranulum* |
| *Ruminococcus* 2 |
| Unclassified *Ruminococcaceae* |
| Unclassified *Enterobacteriaceae* |
| *Akkermansia* |
| *Prevotella* 9* |
| *Bacteroides* |
| *Bifidobacterium* |
| *Ruminococcaceae* UCG-014 |
| Unknown |
| Unclassified *Lachnospiraceae* |
| *Methanobrevibacter* |
| *Phascolarctobacterium* |
| *Acidaminococcus* * |
| *Treponema* 2 |
| *Parabacteroides* |
| *Christensenellaceae* R-7 group |
| *[Eubacterium]* ruminantium group |
| *Butyrivibrio* * |
| *Streptococcus* |

Asterisk indicates taxa with significantly different relative expression between AL and AH groups (DESeq2 analysis presented in Table S2).

**Table S2.**
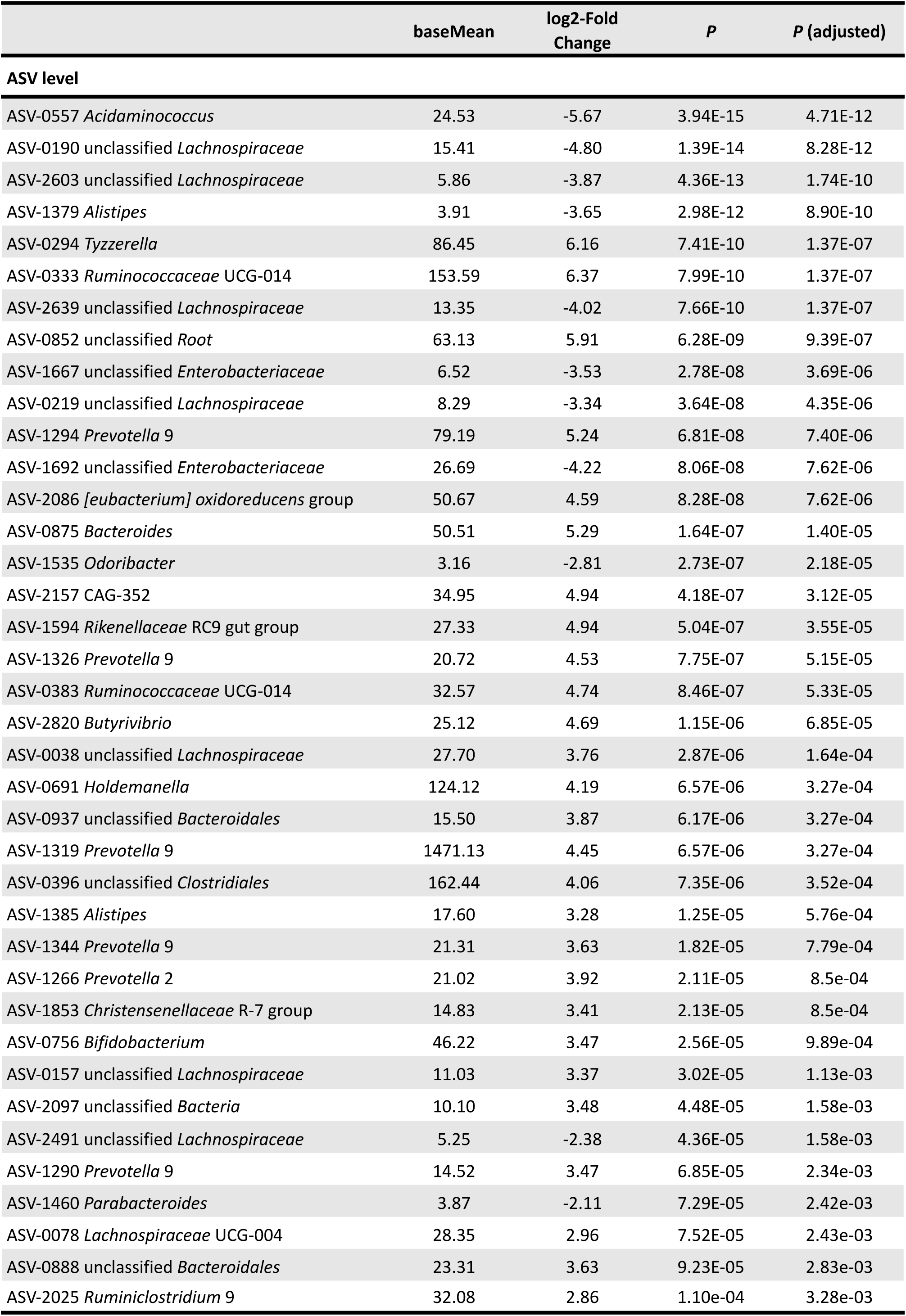
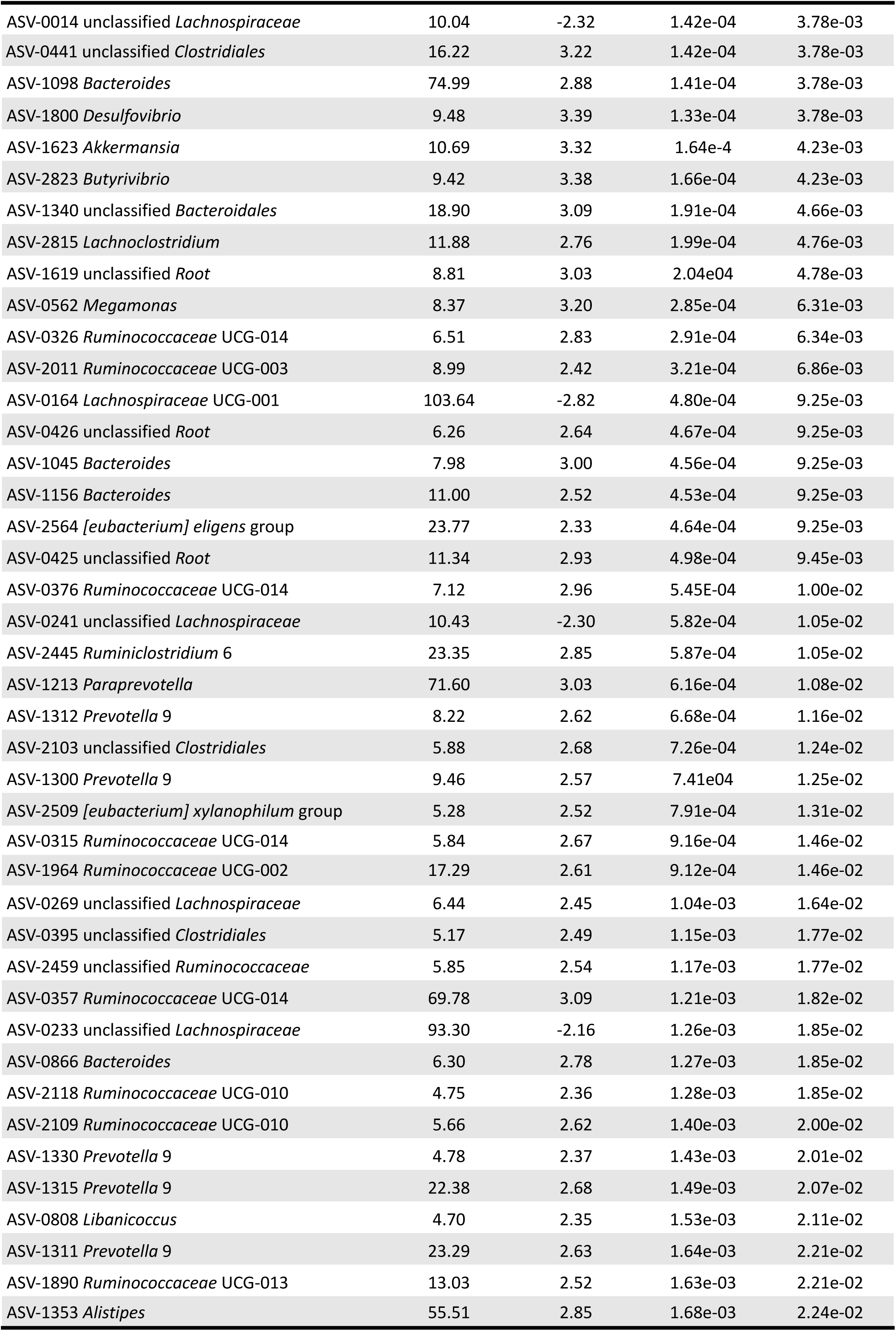
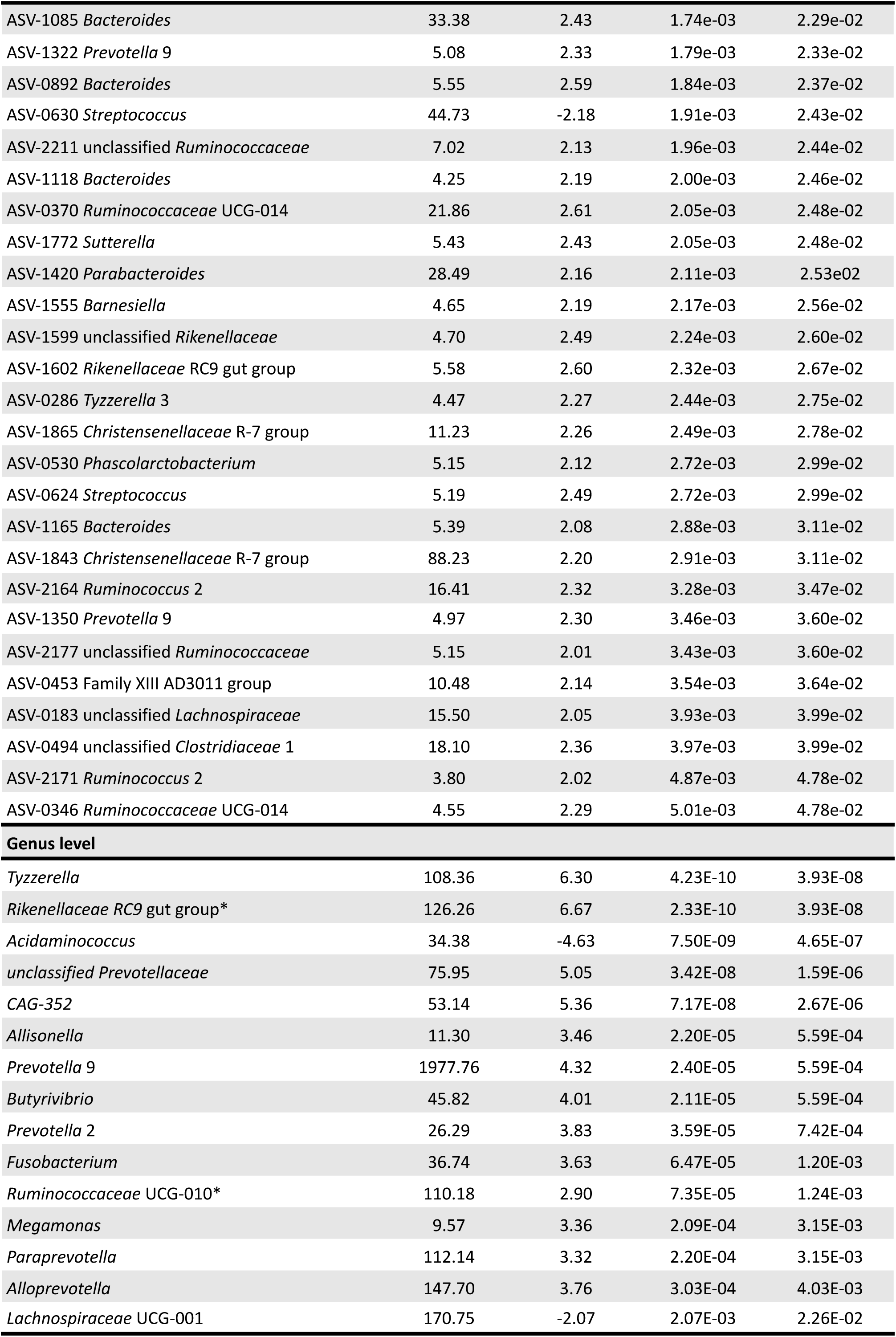
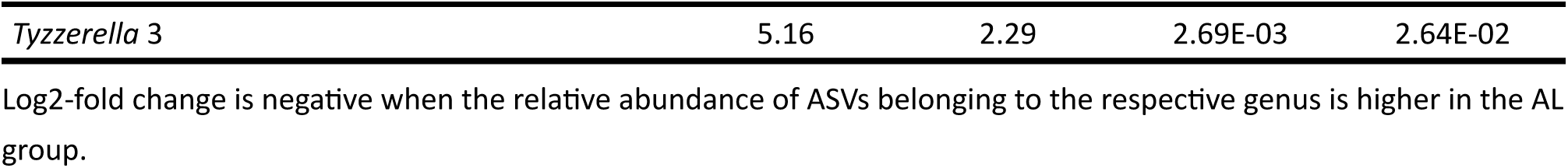
Taxa with significantly different relative abundance in preoperative fecal samples of AL and AH patients; DESeq2 analysis at ASV and genus level.

**Figure S1.**
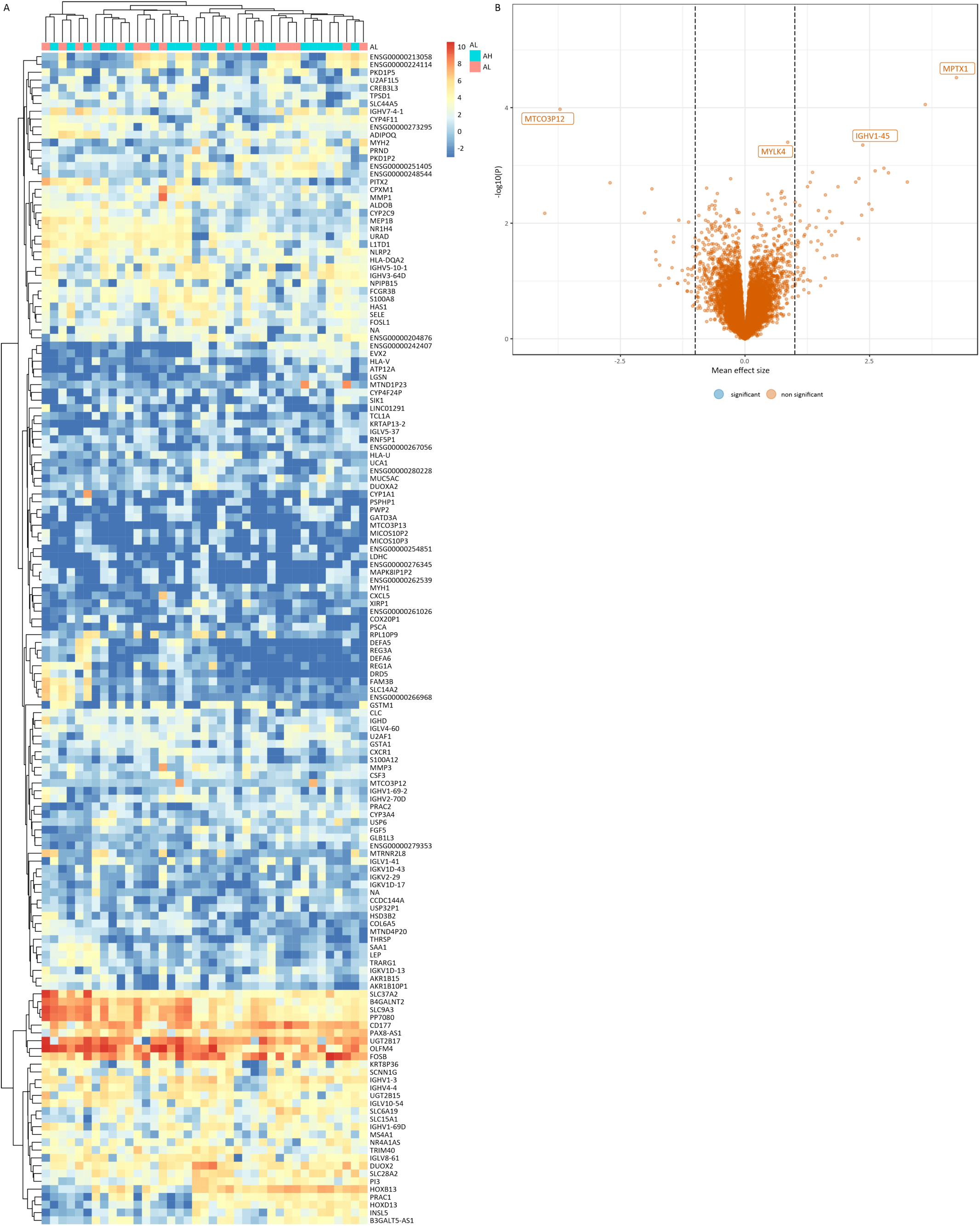
No difference found in the colonic tissue transcriptional profile of patients who undergo colorectal surgery and develop AL. (A) Volcano plot of genes in AL compared to AH patients indicates the absence of significant DEG at surgery. (B) Heatmap representation of the hierarchical clustering analysis of the 150 most variable gene suggests no clear clustering between AL and AH patients.

**Figure S2.**
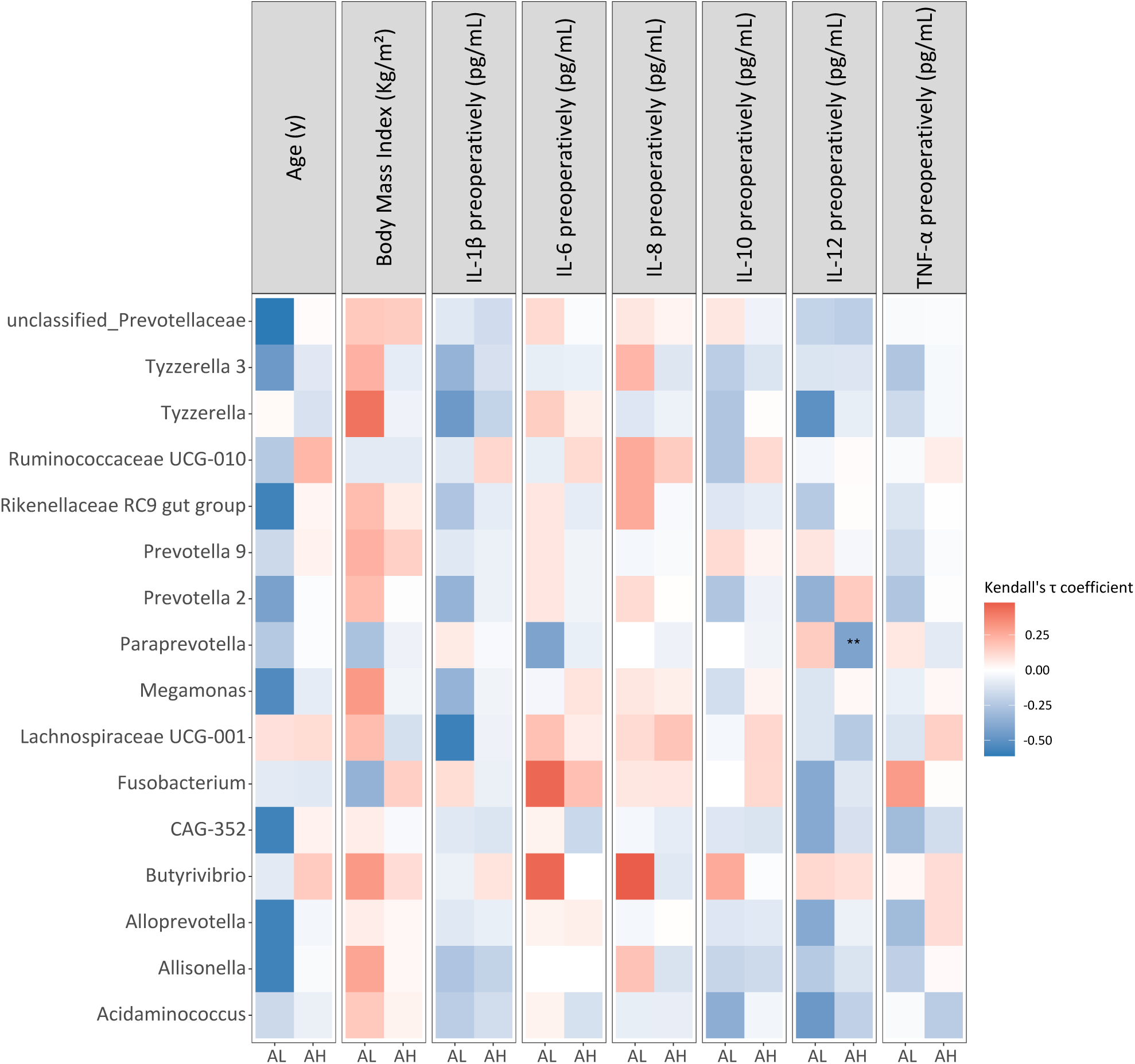
Lack of significant correlation between the microbial discriminatory and preoperative individual components suggests their AL-specific nature. Kendall rank correlations between the relative abundance of the 16 discriminative genera, participants characteristics and preoperative serum cytokines concentrations; Benjamini-Hochberg correction was applied for multiple testing.

